# Time-Restricted Eating Improves Quality of Life, Heart Rate, and Mitochondrial Function in Patients with Postural Orthostatic Tachycardia Syndrome

**DOI:** 10.1101/2025.05.29.25328448

**Authors:** Marissa Dzotsi, Allyssa Strohm, Shweta Varshney, Juan P. Zuniga-Hertz, Ramamurthy Chitteti, Emily Manoogian, Anshika Sethi, Satchidananda Panda, Hemal H. Patel, Taylor A. Doherty, Pam Taub

**Affiliations:** UC San Diego School of Medicine, La Jolla, CA; Salk Institute for Biological Studies, Regulatory Biology, La Jolla, CA; VA San Diego Healthcare Systems, San Diego, CA; UC San Diego, Department of Anesthesiology, La Jolla, CA

**Keywords:** Postural orthostatic tachycardia syndrome, time-restricted eating, pilot study, tachycardia, quality of life, mitochondrial function

## Abstract

Postural orthostatic tachycardia syndrome (POTS) is characterized by an abnormal increase in heart rate upon standing, leading to symptoms such as dizziness, fatigue, and rapid heart rate. Time-restricted eating (TRE), which limits caloric intake to an 8-10 hour daily window, has been shown to decrease inflammation and improve immune, autonomic, and mitochondrial function, as well as cardiometabolic parameters. This single arm pilot study evaluated the effects of TRE on quality of life (QOL), heart rate, and mitochondrial function in 20 participants with POTS (≥30 bpm increase in upright heart rate) and a baseline dietary window of ≥12 hours. Following a 2-week baseline monitoring period, participants underwent a 12-week TRE intervention. Pre- and post-intervention assessments included QOL questionnaires, a 10-minute stand test, and plasma mitochondrial analysis. TRE significantly reduced heart rate increase upon standing (mean decrease: 11 bpm, p < 0.001), improved QOL metrics including POTS symptom severity (p < 0.0001), physical functioning (p = 0.02), and energy/fatigue (p < 0.01), and increased mitochondrial-derived ATP. These findings suggest TRE as a promising lifestyle intervention to improve QOL, heart rate, and mitochondrial function in POTS patients.

## Introduction

Postural orthostatic tachycardia syndrome (POTS) is a heterogeneous, complex clinical syndrome that impacts multiple organ systems ^1^. This syndrome, characterized by an abnormal increase in heart rate upon standing without a corresponding drop in blood pressure, has gained increasing recognition, particularly due to its association with long COVID/post-acute sequelae of SARS-CoV-2 infection (PASC)^2,3^. It is estimated that up to 70% of patients with PASC have POTS, with clinicians worldwide seeing an increasing number of POTS patients ^4^. Despite this increasing prevalence of POTS due to long COVID/PASC, there is very little mechanistic insight into POTS and limited pharmacological options. There are also very few clinical trials examining the efficacy of lifestyle interventions for POTS ^5,6^.

Current diagnostic criteria define POTS as a heart rate increase of at least 30 beats per minute within 10 minutes of standing (or to at least 120 beats per minute), often accompanied by symptoms such as orthostatic intolerance, fatigue, “brain fog,” gastrointestinal dysmotility, and autonomic dysfunction ^7,8^. Similar to other chronic diseases, including chronic obstructive pulmonary disease and congestive heart failure, POTS imposes significant limitations on quality of life and daily functioning^9^. The mechanisms underlying POTS are poorly understood and are thought to be due to a combination of immune, autonomic dysfunction, hypovolemia, and hyperadrenergic state^10^.

Circadian rhythm dysfunction (CRD) is starting to be recognized as a common feature in conditions like POTS and myalgic encephalomyelitis (ME)/chronic fatigue syndrome (CFS) ^11^. CRD disrupts the natural 24-hour biological cycles that regulate sleep-wake cycles, hormone secretion, and metabolic activity. This disruption can lead to mitochondrial dysfunction, impaired immune responses, and dysregulation of the autonomic nervous system, exacerbating symptoms in affected individuals ^12^.

Emerging research suggests that interventions targeting circadian rhythm disturbances, such as time-restricted eating (TRE), may hold promise in managing symptoms associated with POTS. TRE involves restricting daily food intake to an 8-10 hour window, aligning meal times with the body’s circadian clock. Both animal and human studies have shown that TRE can improve circadian rhythm alignment, mitochondrial, immune, and autonomic function, reduce inflammation, and improve overall cardiometabolic health ^13–16^.

We hypothesized that TRE, by optimizing circadian rhythms and improving autonomic/immune function, could address and improve underlying mechanisms contributing to POTS symptoms and ultimately improve quality of life for affected individuals. This proof-of-concept pilot study aims to evaluate the potential TRE as a lifestyle intervention for POTS.

## Methods

This clinical study was approved by the University of California, San Diego Institutional Review Board and registered on ClinicalTrials.gov (<u>NCT05409651</u>). All patients provided informed written consent.

### Study Design

Following recruitment by the study team, and a 2-week baseline period, eligible patients with an eating window of ≥12-hours were entered into a 12-week TRE intervention, during which they adhered to an 8-10-hour eating window (Figure 1A).

**Figure 1.**
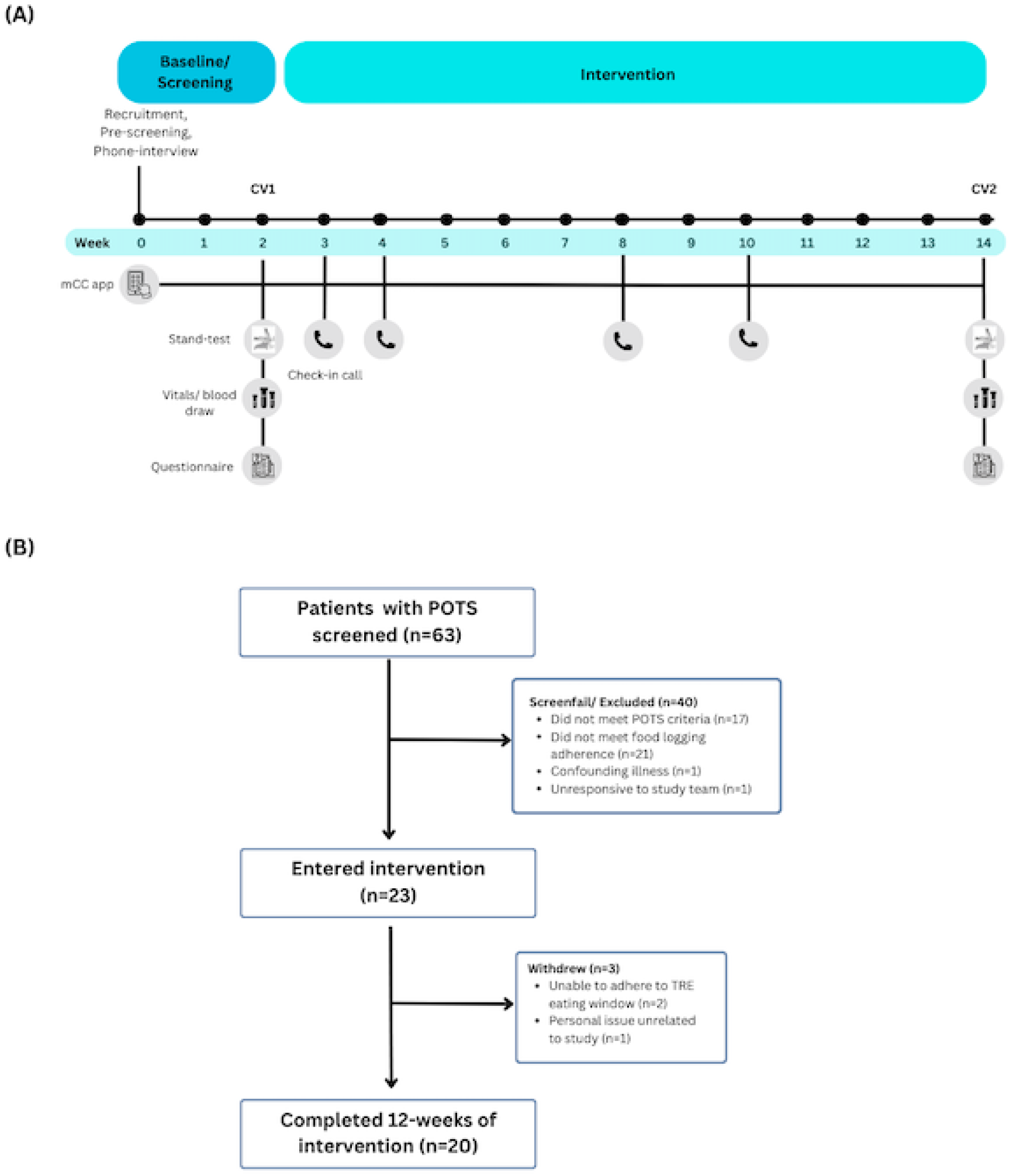
Study timeline and consort diagram. **(A)** The 14-week study consisted of a 2-week baseline period followed by 12-weeks of TRE intervention, with 2 clinic visits (CV). mCC app = myCircadianClock (Gill and Panda, 2015); stand-test = 10-minute POTS orthostatic diagnostic test; questionnaire = SF-36 Health Survey and BDI-II Depression Survey. **(B)** 63 patients were screened for the study, 23 entered intervention, and 20 completed the study and were included in the analysis.

### Patient Selection

Participants were screened and recruited from the cardiovascular and internal medicine clinics at the University of California, San Diego, and from www.clinicaltrials.gov. Patients ages 18-70 years with a self-reported dietary intake of ≥12-hour window, regular daytime schedule of activity (no shift workers) before study enrollment, and a formal diagnosis of POTS with chronic symptoms that have lasted for longer than six months, were enrolled. Pregnant and/or breastfeeding women and patients with other disorders, medications, or functional states that are known to predispose to orthostatic tachycardia, were excluded (Supplemental Material 1). All patients were taking medications for POTS, including beta blockers, midodrine, ivabradine, and fludrocortisone. On average, they were diagnosed with POTS 3.5 years before enrolling in the study, with 3 participants having a post-COVID POTS diagnoses.

### myCircadianClock (mCC) App

Participants who met preliminary eligibility entered a 2-week baseline phase of screening where they were instructed to utilize the mCC app to record all ingestion events. The study team tracked patient eating patterns to confirm ≥12 hour eating windows and determine individual 8-10 hour TRE intervention windows. The length of this TRE interval was at least 4 hours shorter than their average eating window during their pre-screening, with a minimum eating window of 8 hours (from a 12-hour eating window baseline) and a maximum of 10 hours (from a baseline of 14-hours or greater). The interval was entered in the app, so they visualize their daily eating pattern and consume all meals within the designated interval. Participants continued to utilize the mCC app throughout the 12 weeks of intervention and received study-related nudges and reminders from the study team to encourage adherence to the intervention (Supplemental Table 1).

**Table 1.** Baseline Characteristics.

| Table 1. Baseline Characteristics |  |
| --- | --- |
| Age | 36.30 (11.80) |
| BMI | 26.53 (5.61) |
| Sex |  |
| Female | 18 (90.00%) |
| Male | 2 (10.00%) |
| Race |  |
| White | 18 (90.00%) |
| Black | 1 (5.00%) |
| Asian | 1 (5.00%) |
| Ethnicity |  |
| Hispanic | 2 (10.00%) |
| Non-Hispanic | 17 (85.00%) |
| Unknown | 1 (5.00%) |
| POTS Medication |  |
| Fludrocortisone | 2 (10.00%) |
| Ivabradine | 13 (65.00%) |
| Midodrine | 4 (20.00%) |
| Propranolol | 4 (20.00%) |
| Heart Rate, beats/min |  |
| Supine | 70.45 (13.23) |
| Standing | 114.55 (14.44) |
| Systolic Blood Pressure, mm HG |  |
| Supine | 118.00 (12.56) |
| Standing | 124.05 (11.76) |
| Diastolic Blood Pressure, mm HG |  |
| Supine | 74.30 (11.24) |
| Standing | 84.85 (12.21) |
| N= 20. Values are presented as mean (SD) for continuous variables and n (%) for categorical variables. |  |

The smartphone mCC serves as an electronic food, activity, and sleep diary. If a participant fails to log any food data for more than 1 day, the dashboard flags the participant and sends an alert to the research coordinator. The coordinators will login to the dashboard at least twice weekly to monitor food intake data, and follow up with flagged participants as necessary. If any participant faces difficulty logging data, or has questions about any of the features of the app, they can contact the study coordinator through the feedback feature of the app. Their questions are encrypted and delivered to a HIPAA compliant email server specifically set up for this study.

### Intervention

Following a 2-week baseline period, participants whose initial eating windows met study criteria were invited to the clinic for their first visit. Patients were instructed to hold their medications for POTS for 2 days before Visit 1. During Visit 1, a 10-minute active standing test was conducted to confirm the diagnosis of POTS and establish baseline orthostatic values without medications ^17^. Participants failing to meet POTS criteria (30-point increase in heart rate) during this test were excluded from further study procedures. After the 10-minute active standing test, a fasted blood draw was performed, and participants completed general health and sleep questionnaires. Screening for depression utilized the Beck Depression Inventory-II (BDI-II), with scores ≥19, without stable medication or therapy, resulting in exclusion. Quality of life measurements were assessed using the Malmö POTS Symptom Score Survey (MAPS) and the General Health Questionnaire Short Form-36 (SF-36).

Upon completion of Visit 1, data was reviewed, and eligible participants worked with a research coordinator to select an 8-10 hour eating window tailored to fit their lifestyle, based on pre-screening eating patterns. Subsequently, participants entered a 12-week intervention period during which dietary intake was documented using the mCC app. Participants were instructed to hydrate well during the fasting period and not to make any other changes to their diet. Periodic phone contacts reinforced nutritional practices, supplemented by daily push notifications on their phones.

After the 12-week intervention, participants returned to the clinic for Visit 2, replicating procedures from Visit 1, including the standing test (with medications held for 2 days before visit similar to visit 1), fasted blood draw, and health questionnaires.

### Outcomes

The primary outcome was a change in self-reported quality of life (QOL), evaluated using two instruments: the 36-Item Short Form Health Survey questionnaire (SF-36) and the Malmö POTS Symptom Score (MAPS) survey. The SF-36 is widely used to assess health-related quality of life, providing insights into patients’ physical and mental well-being ^18^.

The MAPS survey was specifically developed to assess the symptom burden experienced by POTS patients. It utilizes a semiquantitative system to evaluate the severity of 12 commonly reported symptoms: five cardiac symptoms (palpitations, dizziness, presyncope, dyspnea, and chest pain) and seven non-cardiac symptoms (gastrointestinal symptoms, insomnia, concentration difficulties, headache, myalgia, nausea, and fatigue)^19^. Unlike the SF-36, which offers a broader evaluation of health-related QOL, the MAPS survey focuses specifically on symptoms relevant to POTS, providing detailed and condition-specific insights into patient experiences.

A secondary outcome of this study was the change in orthostatic heart rate (HR) when patients transitioned from a supine to an upright position. This was assessed using a 10-minute standing test, during which the difference between the supine HR and the highest HR recorded within the 10-minute standing period was measured.

To analyze the exploratory outcome of mitochondrial function, glycolytic and mitochondrial metabolism profiling was performed using established techniques^20^. To analyze the metabolic effects of plasma samples, C2C12 cells, mouse myoblasts (ATCC, CRL-1772™) were used. C2C12 cells were maintained in culture with DMEM (ATCC, 30-2002™) supplemented with 10% fetal bovine serum (Gibco, A31604-01), 37°C, 5% CO2, and controlled humidity. To perform the metabolic profiling assays, cells were resuspended with trypsin 0.25% (Gibco, 25200-056), centrifuged at 1,000RPM for 5min, and washed with PBS1X (Gibco, 10010-023). The cells were seeded in Agilent Seahorse 96-well plates (Agilent, 103774-100) at a density of 10,000 cells per well and maintained in incubation for 24h before the assay. All cells used for the metabolic profiling assays were at or before passage 5.

A glycolytic and mitochondrial metabolism profile was performed to analyze the plasma samples’ potential metabolic effects. C2C12 cells were treated with plasma samples (plasma 1%) prepared in Agilent Seahorse Assay Media (XF DMEM 103575-100, HEPES 5mM) supplemented with Agilent glucose (103577-100, final concentration 10mM), glutamine (103579-100, final concentration 2mM, and pyruvate (103578-100, final concentration 1mM). The cells were washed twice in supplemented Seahorse Assay Media, treated with plasma solutions (100mL final volume per well), and incubated for 1h in a CO2-free incubator as part of the standard Agilent Seahorse degassing procedure. After 1h, the media was removed and replaced with fresh Agilent Seahorse Assay Media (180mL final volume per well) and a Mitochondrial Stress assay (Agilent Cell Mito Stress Test Kit, 103015-100) and a Real-time ATP Rate Assay (Agilent Real-Time ATP Rate Assay Kit, 103592-100) were performed in an Agilent Seahorse XF Pro. Both assays were run in parallel in two Seahorse XF Pro systems for data reliability. The final well concentrations of the Mitochondrial Stress assay used were oligomycin at 1.5mM, FCCP at 2.0mM, and rotenone/antimycin A at 0.5mM. For the Real-time ATP Rate Assay, the final well concentrations of the compounds used were oligomycin at 1.5mM and rotenone/antimycin A at 0.5mM. In both assays, during the Rotenone/Antimycin A injection Hoechst (ThermoScientific, 33342) was added. All data was normalized by cell count per well and presented as Oxygen Consumption Rate (OCR) (pmol O2/min/1,000 cells). ATP data is presented as ATP production rate (pmol/min/1,000 cells).

### Statistical methods

Descriptive statistics and exploratory graphing techniques, including frequencies, means, standard deviations (SDs), box and whisker plots, stem and leaf diagrams, and scatter plots, were employed to assess the data for normality, skewness, and outliers. Additionally, z-scores for skewness and kurtosis were calculated for all outcomes. Final analyses for heart rate and quality of life were conducted using both the original data (non-log-transformed) and nonparametric methods, yielding similar results. All reported analyses utilized the original non-log-transformed data format.

The comparability of baseline and post-intervention values was tested using analyses of variance (ANOVAs) for continuous variables and chi-square analyses for dichotomous variables. Treatment effect estimates, along with corresponding p-values, were derived from within-patient comparisons. All statistical tests were two-tailed. All available data was included in the analyses, with no patients excluded due to missing data. The SF-36 was scored according to RAND instructions ^21^. Differences were considered statistically significant if a p-value of 0.05 or less was obtained using R version 4.1.1.

## Results

### Patient Demographics

Baseline characteristics are shown in Table 1. In total, 63 patients were screened, 40 failed screening/ were excluded (e.g. did not meet POTS criteria, did not meet food logging adherence, confounding illness). Of the 23 that entered the 12-week intervention, 3 withdrew (unable to adhere to TRE eating window and personal issues unrelated to the study), and 20 completed the study (average age 36.3 ± 11.8 years, n = 18 women) and were included in the final data analysis (Figure 1B).

The majority of participants who completed the study were White (90%), Non-Hispanic (85%), and predominantly female (90%). All participants met formal diagnostic criteria for POTS and maintained stable doses of pharmacological therapy throughout the study period.

### Primary Outcome: Quality of Life

The SF-36 survey provided a comprehensive assessment of various aspects related to QOL. Analysis of baseline and post-intervention data revealed significant improvements in several domains. Specifically, participants reported enhanced physical function (Delta = 9.00 (1.98 to 16.02); p = 0.02) and improved levels of energy/fatigue (Delta = 11.25 (5.09 to 17.41); p<0.01) following the 12-week intervention (Table 2A).

**Table 2.** The Effect of TRE on Quality of Life Surveys.

| Table 2. The Effect of TRE on Quality of Life Surveys |  |  |  |  |
| --- | --- | --- | --- | --- |
| Outcomes | Baseline | Post Intervention | Delta (95% CI) | p-value |
| **A. Short-Form 36 (SF-36).** Higher scores represent a higher quality of life. |  |  |  |  |
| Physical Functioning | 52.75 (20.87) | 61.75 (16.24) | 9.00 (1.98 to 16.02) | 0.02* |
| Role Limitations Due to Physical Health | 21.25 (24.70) | 31.25 (31.28) | 10.00 (-8.75 to 28.75) | 0.28 |
| Role Limitations Due to Emotional Problems | 65.00<br>(36.64) | 65.00<br>(42.54) | 0.00 (-25.31 to 25.31) | 1.00 |
| Energy/Fatigue | 19.75<br>(12.40) | 31.00<br>(15.78) | 11.25 (5.09 to 17.41) | <0.01** |
| Emotion Well-Being | 56.00<br>(15.57) | 59.80<br>(16.74) | 3.80 (-2.09 to 2.09) | 0.31 |
| Social Functioning | 56.88<br>(19.65) | 65.00<br>(21.31) | 8.12 (-2.14 to 18.39) | 0.11 |
| Pain | 69.38<br>(20.28) | 67.63<br>(19.84) | -1.75 (-8.47 to 4.97) | 0.59 |
| General Health | 36.25<br>(15.46) | 37.25<br>(16.10) | 1.00 (-4.18 to 6.18) | 0.69 |
| Health Change | 57.50<br>(27.02) | 57.50<br>(31.52) | 0.00 (-10.74 to 10.74) | 1.00 |
| **B. Malmo POTS Symptom Survey (MAPS Survey).** Lower scores represent a lower severity of symptoms. |  |  |  |  |
| Dizziness | 5.25 (2.47) | 3.45 (2.46) | -1.80 (-2.73 to -0.87) | <0.01** |
| Feeling Faint | 3.40 (2.89) | 2.75 (2.73) | -0.65 (-1.76 to 0.46) | 0.23 |
| Palpitations | 7.15 (2.62) | 5.40 (2.98) | -1.75 (-2.78 to -0.72) | <0.01** |
| Difficulty Breathing | 3.95 (2.96) | 2.60 (2.06) | -1.35 (-2.39 to -0.31) | 0.01* |
| Chest Pain | 2.70 (3.42) | 2.10 (2.85) | -0.60 (-1.61 to 0.41) | 0.23 |
| Headache | 4.30 (3.37) | 3.45 (3.25) | -0.85 (-2.15 to 0.45) | 0.19 |
| Concentration | 5.70 (2.85) | 4.35 (2.94) | -1.35 (-2.75 to 0.05) | 0.06 |
| Muscle Pain | 4.15 (3.00) | 3.45 (3.07) | -0.70 (-2.13 to 0.73) | 0.32 |
| Nausea | 3.60 (3.12) | 2.05 (2.37) | -1.55 (-2.62 to -0.48) | <0.01** |
| Gastrointestinal Issues | 6.25 (3.01) | 3.90 (3.37) | -2.35 (-3.82 to -0.88) | <0.01** |
| Exhaustion | 8.35 (1.39) | 5.00 (3.28) | -3.35 (-4.69 to -2.01) | <0.001*<br>** |
| Insomnia | 4.95 (3.44) | 2.15 (1.98) | -2.80 (-4.51 to -1.09) | <0.01** |
| Total Severity Score | 59.75 (19.08) | 40.65 (20.04) | -19.1 (-26.50 to -11.70) | <0.001*<br>** |
| N=20. **2A.** TRE significantly improved reported SF-36 survey scores of physical functioning (p =0.02) and energy/ fatigue (p<0.01). **2B.** The severity of reported POTS symptoms from the MAPS survey showed significant improvement following TRE.<br>Data is shown as Mean (SD) and Mean (95% Confidence Interval) for delta values. (*) = p<0.05, (**) = p<0.01, (***) =p<0.001. |  |  |  |  |

Analysis of the MAPS survey revealed significant improvements across multiple common POTS symptoms following the 12-week intervention (Table 2B). Participants reported notable reductions in symptoms such as dizziness, palpitations, difficulty breathing, nausea, gastrointestinal issues (GI), exhaustion, and insomnia. The average overall severity score decreased by approximately 19 points (p < 0.001). Notably, the average severity score post-intervention was 40.65, indicating a significant improvement from baseline. The optimal cut-point value of MAPS to differentiate POTS from healthy controls is a score ≥42, underscoring the clinical significance of these improvements ^19^.

### Secondary Outcome: Orthostatic Heart Rate

Upon analyzing the secondary outcome of orthostatic heart rate, we observed notable changes in participants’ cardiovascular responses throughout the study period (Table 3). At baseline, participants exhibited an average supine heart rate of 70.45 (±13.23) beats per minute (bpm), which increased significantly to an average of 114.55 (±14.44) bpm upon assuming an upright posture. Following the 12-week TRE intervention, a marked improvement in orthostatic heart rate was observed (p < 0.0001). Specifically, the increase in orthostatic heart rate between baseline and post-intervention decreased by approximately 11 bpm (44.1 bpm vs. 33.25 bpm). Post-intervention, participants’ supine heart rate averaged 71.15 (±12.97) bpm, with an average standing heart rate of 104.4 (±16.41) bpm.

**Table 3.** The Effect of TRE on Heart Rate.

|  | Baseline | Post Intervention | Delta (95% CI) | p-value |
| --- | --- | --- | --- | --- |
| Supine Heart Rate, (beats/min) | 70.45 (13.23) | 71.15 (12.97) | 0.70 (-5.72 to 4.32) | 0.77 |
| Standing Heart Rate, (beats/min) | 114.55 (14.44) | 104.4 (16.41) | -10.15 (4.37 to 15.93) | <0.001** |
| Delta Heart Rate, Standing vs. Supine (beats/min) | 44.1 (14.75) | 33.25 (12.84) | -10.85 (7.56 to 14.14) | <0.0001*** |
| N=20. TRE led to a significant decrease in standing heart rate (p<0.001) and delta heart rate (p<0.0001). |  |  |  |  |
| Data is shown as Mean (SD) and Mean (95% Confidence Interval) for delta values. (*) = p<0.01, (**) = p<0.001, (***) = p<0.0001. |  |  |  |  |

### Exploratory Outcome: Mitochondrial Function

Agilent Seahorse Metabolic Flux assessments revealed no significant changes in mitochondrial stress test outcomes between Pre- and Post-TRE plasma-treated cells (Figure 2A and 2B). However, under physiological conditions, there was a significant increase in mitochondrial-derived ATP synthesis in cells treated with Post-TRE plasma compared to Pre-TRE plasma (Figure 2C). These findings suggest that factors elevated in Post-TRE plasma may enhance cellular metabolism and energy production without affecting the mitochondrial stress response.

**Figure 2.**
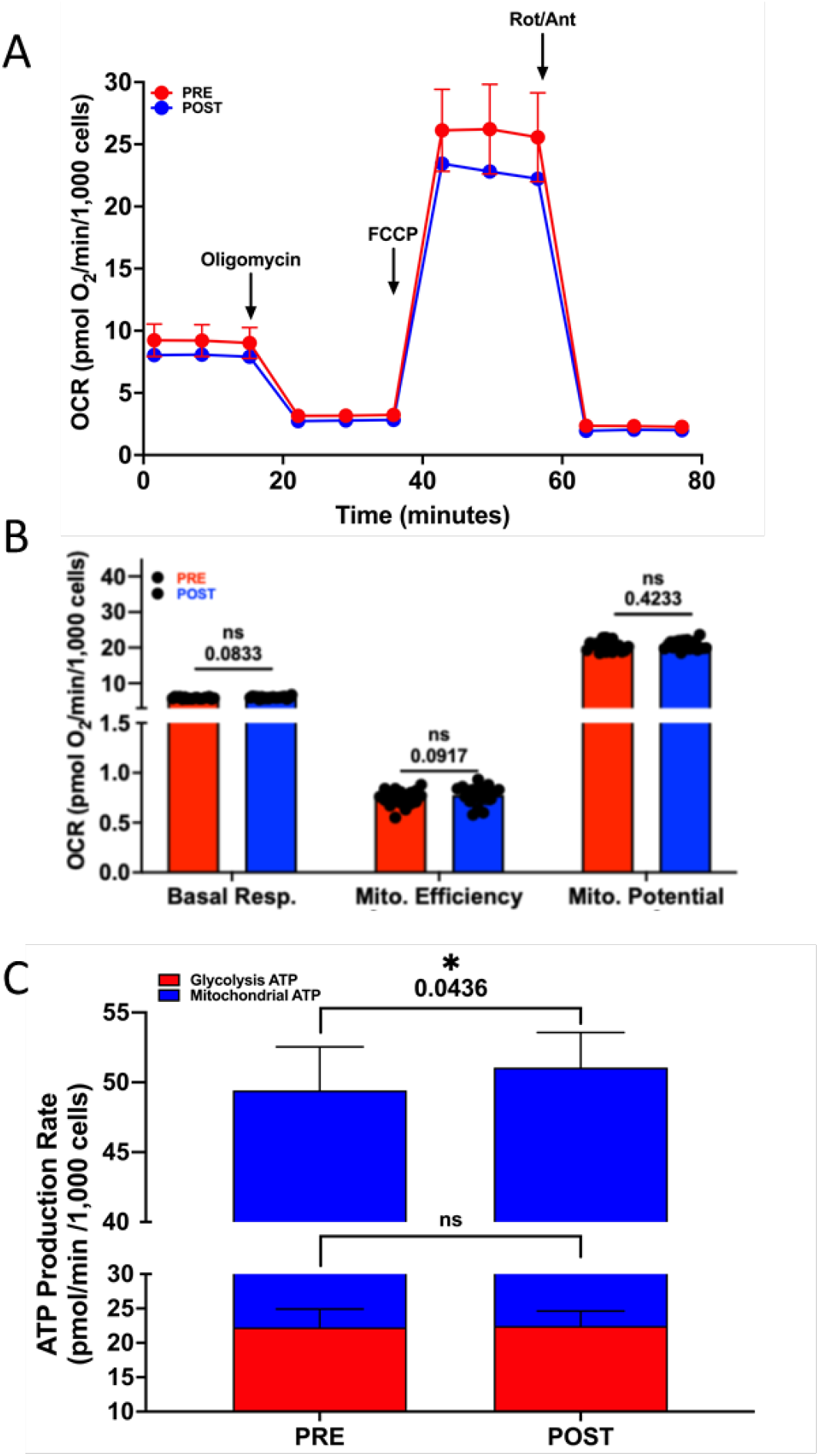
Pre and post TRE plasma-mediated alteration of mitochondrial respiration in skeletal muscle cells. We utilized the Agilent Seahorse Metabolic Flux system to assess mitochondrial stress (**A** and **B**) and metabolic profile (**C**) by monitoring oxygen consumption rates (OCR) via adoptive transfer of plasma to cultured C2C12 cells. C2C12 cells were incubated with plasma at 1% final concentration for one hour before initiating the mitochondrial stress energy generation protocol. **A**. Averaged values representing tracing of the Seahorse protocol. We assessed baseline respiration up until the addition of oligomycin (ATPase inhibitor) followed by FCCP (uncoupler) and finally rotenone/antimycin A (Rot/Ant, complex I and III inhibitors, respectively). **B**. Mitochondrial functional parameters were analyzed for all samples. We saw no change in mitochondrial stress parameters. **C**. Metabolic profiling revealed a significant increase (p<0.05) in mitochondrial-derived ATP in post-TRE plasma, suggesting that factors in the post-TRE plasma can potential increase mitochondrial function. Groups were n=20.

## Discussion

POTS presents substantial challenges in management due to its complex symptomatology and limited treatment options. This study explored the impact of a 12-week time-restricted eating intervention on POTS patients and observed significant improvements across various outcomes. Quality of life assessments using the SF-36 and Malmö POTS Symptom Score surveys indicated significant enhancements in physical function (p = 0.02), energy/fatigue (p < 0.01), and reductions in symptom severity encompassing domains such as dizziness, palpitations, and gastrointestinal issues (p < 0.001). Participants demonstrated a notable decrease in orthostatic heart rate, averaging an 11 beats per minute reduction (p < 0.0001) after the TRE intervention. Importantly, 20 out of 23 participants completed the intervention, underscoring high adherence to the TRE protocol within our study cohort, which predominantly comprised middle-aged women of White and Non-Hispanic ethnicity. There were no adverse events associated with TRE in this population.

Time-restricted eating emerges as a pragmatic intervention for POTS patients for several reasons. Unlike more restrictive dietary approaches, TRE accommodates personal food choices and does not necessitate calorie reduction or increased physical activity, which can be challenging for POTS patients ^22^. It requires no higher level of health literacy, is culturally acceptable, intuitive to follow, and can complement pharmacotherapy. Our study specifically implemented a 10-hour TRE protocol, which proved safe, feasible, and associated with high adherence, without adverse effects ^23^. TRE has been shown in both animal and clinical trial to optimize circadian rhythms, promotes a safe state of ketosis, enhances mitochondrial function, reduces inflammation, and improves autonomic function—and has potential to address underlying pathophysiological derangements in POTS resulting in improvement in the quality of life for POTS patients ^23 24^.

In line with these broader implications, our exploratory analysis of mitochondrial function revealed that cells treated with Post-TRE plasma demonstrated a significant increase in mitochondrial-derived ATP synthesis under physiological conditions compared to Pre-TRE plasma-treated cells, suggesting that TRE may improve mitochondrial function in patients with POTS. Studies have shown the impactful relationship between mitochondrial dysfunction and POTS is seen in long COVID/ PASC patients, whose mitochondrial damage in the peripheral and central nervous systems may disrupt the coordination of autonomic responses, exacerbating symptoms such as fatigue and tachycardia ^25^. Improved mitochondrial function could contribute to the observed reductions in fatigue and improvements in physical function and overall well-being.

Addressing orthostatic heart rate is pivotal in managing POTS due to its diagnostic significance and therapeutic implications. Monitoring and managing orthostatic heart rate abnormalities are essential for tailoring effective treatment strategies that alleviate symptoms and enhance overall well-being in individuals with POTS ^26,27^. The observed 11 bpm reduction in orthostatic heart rate post-TRE (44.1 bpm vs. 33.25 bpm, p < 0.0001) suggests enhanced autonomic function and improved cardiovascular stability, potentially alleviating symptoms such as lightheadedness, dizziness, and fatigue associated with POTS. This 11 bpm reduction is comparable to what is seen with pharmacologic interventions for POTS such as beta blockers ^28^.

Quality of life outcomes further underscored the intervention’s efficacy, revealing significant improvements in common POTS symptoms. Participants reported marked enhancements in physical function, increased energy levels, and reduced symptom severity across multiple domains, including dizziness, palpitations, and gastrointestinal issues (p < 0.001). These findings are consistent with previous research linking improvements in orthostatic heart rate to enhanced symptom management and overall quality of life in POTS patients ^29,30^. Notably, symptoms such as gastrointestinal issues are prevalent among POTS patients with connective tissue diseases and autonomic neuropathy ^31^, often co-occurring with conditions like Ehlers-Danlos syndrome and functional GI disorders ^32^. Addressing these multifaceted symptoms by targeting mitochondrial function and autonomic dysfunction and improving the hemodynamic aspects of POTS, TRE not only mitigates physiological dysregulation but also translates into substantial improvements in daily functioning and overall well-being for individuals living with POTS. Importantly, the majority of POTS patients in our study were severe enough to be treated chronically with pharmacotherapy and TRE was able to further improve symptoms beyond what medications could provide.

## Limitations

This is a single-center clinical trial with a small sample size. However, the sample size is comparable to other published clinical studies in POTS. This is also an open-label study as it is difficult to have blinding with a dietary intervention such as TRE. While our study showed short-term benefits over the 12-week period, the durability of these improvements beyond the intervention phase requires further investigation. Larger studies with longer duration of TRE are warranted to confirm these findings.

## Conclusion

Interventions aimed at optimizing orthostatic heart rate are pivotal in tailoring effective treatment approaches for POTS patients. Our findings highlight TRE as a promising addition to existing pharmacological strategies, offering a non-pharmacological avenue to modulate cardiovascular responses and improve overall well-being in individuals with POTS.

## Data Availability

The data underlying this article will be shared upon reasonable request to the corresponding author.

## Acknowledgments

This work was supported by a research grant from Dysautonomia International to PT. Research in SP lab is supported by Clara Wu and Joe Tsai Foundation and Wu Tsai Human Performance Alliance. HHP is supported by a Veterans Administration Research Career Scientist Award (BX5005229). ENM is supported by the Larry L Hillblom Foundation and the National Institute of Diabetes and Digestive And Kidney Diseases of the National Institutes of Health under Award Number R01DK139356. The content is solely the responsibility of the authors and does not necessarily represent the official views of the National Institutes of Health.

## Notes

### Competing Interest Statement

The authors have declared no competing interest.

### Clinical Trial

NCT05409651

### Funding Statement

This study was funded by Dysautonomia International.

### Author Declarations

Ethics committee/ IRB of the University of California, San Diego gave ethical approval for this work.

