## Supplemental Materials for "Time-Restricted Eating Improves Quality of Life, Heart Rate, and Mitochondrial Function in Patients with Postural Orthostatic Tachycardia Syndrome"

Supplemental Material 1: Inclusion/Exclusion Criteria

Inclusion Criteria:

1. Age: 18-70 years old
2. POTS, as defined by the presence of any of the following criteria:
   - For patients age 20 or older, increase in heart rate ≥ 30 bpm within ten minutes of upright posture (tilt test or standing) from a supine position (For patients age 18-19, heart rate increase must be >40 bpm)
   - Associated with related symptoms that are worse with upright posture and that improve with recumbency
   - Chronic symptoms that have lasted for longer than six months
   - In the absence of other disorders, medications, or functional states that are known to predispose to orthostatic tachycardia
3. Baseline eating period > 12-hour window

Exclusion Criteria:

1. Taking insulin within the last 6 months.
2. Manifest diabetes, defined as HbA1c > 7.0% given a 0.3% margin of error in lab readings, or diagnosis of diabetes.
3. Known inflammatory and/or rheumatologic disease.
4. Active tobacco abuse or illicit drug use or history of treatment for alcohol abuse.
5. Pregnant or breast-feeding women.
6. Shift workers with variable (e.g. nocturnal) hours.
7. Caregivers for dependents requiring frequent nocturnal care/sleep interruptions.
8. Planned travel to a time zone with greater than a 3-hour difference during study period.
9. History of a major adverse cardiovascular event within the past 1 year (acute coronary syndrome (ACS), percutaneous coronary intervention, coronary artery bypass graft surgery, hospitalization for congestive heart failure, stroke/transient ischemic attack (TIA)).
10. Uncontrolled arrhythmia (i.e. rate-controlled atrial fibrillation/atrial flutter are not exclusion criteria).
11. History of thyroid disease requiring dose titration of thyroid replacement medication(s) within the past 3 months (i.e. hypothyroidism on a stable dose of thyroid replacement therapy is not an exclusion).
12. History of adrenal disease.
13. History of malignancy undergoing active treatment, except non-melanoma skin cancer.
14. Known history of type I diabetes.
15. History of eating disorders.
16. History of cirrhosis.
17. History of stage 4 or 5 chronic kidney disease or requiring dialysis.
18. History of HIV/AIDS.
19. Currently enrolled in a weight-loss or weight-management program.
20. On a special or prescribed diet for other reasons (e.g. Celiac disease).
21. Currently taking any medication that is meant for, or has a known effect on, appetite.
22. Any history of surgical intervention for weight management.
23. Uncontrolled psychiatric disorder (including history of hospitalization for psychiatric illness).
24. A score of >16 on the Epworth Sleepiness Scale (ESS).
25. Depression determined by the Beck Depression Inventory (BDI-II) (unless previously diagnosed and well-controlled)
26. Failure to use the smartphone app for documentation (defined as <2 meals/day for ≥3 days during baseline).

| **Supplemental Table 1. TRE Eating Window and Adherence** | | | |
| --- | --- | --- | --- |
| **Study Phase** | **Baseline** | **Intervention (Last 2 weeks)** | **Intervention (0-12 weeks)** |
| Duration (total days) | 14.55 (1.00) | 13.80 (0.52) | 85.35 (3.51) |
| Number of Good Logging Days | 13.55 (2.06) | 12.65 (1.93) | 73.85 (8.35) |
| Number of caloric entries | 82.8 (30.1) | 53.25 (17.86) | 364 (114.43) |
| 95% eating window (hours) | 15.19 (2.11) | 10.44 (1.95) | 10.72 (1.98) |
| Change in eating window (hours) | - | -4.75 hours | -4.47 hours |
| Percent of days outside the eating window | - | 12% | 10% |
| N=20. Data is shown as mean (SD) for each time point and mean (95% CI) for changes between time points. The data shown is based on entries on the myCircadianClock app. TRE= time-restricted eating. | | | |
